# An adaptive randomized controlled trial of non-invasive respiratory strategies in acute respiratory failure patients with COVID-19

**DOI:** 10.1101/2021.08.02.21261379

**Authors:** Gavin D Perkins, Chen Ji, Bronwen A Connolly, Keith Couper, Ranjit Lall, J Kenneth Baillie, Judy M Bradley, Paul Dark, Chirag Dave, Anthony De Soyza, Anna V Dennis, Anne Devrell, Sara Fairbairn, Hakim Ghani, Ellen A Gorman, Christopher A Green, Nicholas Hart, Siew Wan Hee, Zoe Kimbley, Shyam Madathil, Nicola McGowan, Benjamin Messer, Jay Naisbitt, Chloe Norman, Dhruv Parekh, Emma M Parkin, Jaimin Patel, Scott E Regan, Clare Ross, Anthony J Rostron, Mohammad Saim, Anita K Simonds, Emma Skilton, Nigel Stallard, Michael Steiner, Rama Vancheeswaran, Joyce Yeung, Daniel F McAuley, On behalf of the Recovery- RS collaborators

## Abstract

**Background:** Both continuous positive airway pressure (CPAP) and high-flow nasal oxygenation (HFNO) have been recommended for acute respiratory failure in COVID-19. However, uncertainty exists regarding effectiveness and safety.

**Methods:** In the Recovery-Respiratory Support multi-center, three-arm, open-label, adaptive, randomized controlled trial, adult hospitalized patients with acute respiratory failure due to COVID-19, deemed suitable for treatment escalation, were randomly assigned to receive CPAP, HFNO, or conventional oxygen therapy. Comparisons were made between each intervention and conventional oxygen therapy. The primary outcome was a composite of tracheal intubation or mortality within 30-days.

**Results:** Over 13-months, 1272 participants were randomized and included in the analysis (380 (29.9%) CPAP; 417 (32.8%) HFNO; 475 (37.3%) conventional oxygen therapy). The need for tracheal intubation or mortality within 30-days was lower in the CPAP group (CPAP 137 of 377 participants (36.3%) vs conventional oxygen therapy 158 of 356 participants (44.4%); unadjusted odds ratio 0.72; 95% CI 0.53 to 0.96, P=0.03). There was no difference between HFNO and conventional oxygen therapy (HFNO 184 of 414 participants (44.4%) vs conventional oxygen therapy 166 of 368 participants (45.1%); unadjusted odds ratio 0.97; 95% CI 0.73 to 1.29, P=0.85).

**Conclusions:** CPAP, compared with conventional oxygen therapy, reduced the composite outcome of intubation or death within 30 days of randomisation in hospitalized adults with acute respiratory failure due to COVID-19. There was no effect observed, compared with conventional oxygen therapy, with the use of HFNO.

(Funded by the UK National Institute for Health Research; ISRCTN 16912075).

## INTRODUCTION

Acute respiratory failure is a key clinical characteristic of COVID-19 pneumonitis, with 76% of hospitalised patients requiring supplemental oxygen and 9% requiring tracheal intubation and invasive mechanical ventilation.^1^ Early in the pandemic, international experiences highlighted the potential risk that intensive care units might become overwhelmed, and high mortality in patients that required invasive mechanical ventilation.^2-4^ This drove an urgent public health need to identify strategies that reduce the need for invasive mechanical ventilation.

In COVID-19 patients with increasing oxygen requirements, non-invasive respiratory strategies, such as continuous positive airway pressure (CPAP) and high-flow nasal oxygen (HFNO), provide a potentially attractive strategy for avoiding invasive mechanical ventilation. In other respiratory diseases, particularly community acquired pneumonia, both CPAP and HFNO may improve clinical outcomes, although those treated with CPAP experience more adverse events.^5,6^ In the context of COVID-19, however, there was concern that these strategies might serve only to delay tracheal intubation due to high failure rates, whilst correspondingly exacerbating lung injury through generation of large tidal volumes.^7-10^ At a wider system level, there is ongoing uncertainty around the risk of nosocomial infection with aerosol generation and risks oxygen shortages due to the high demand placed on hospital oxygen delivery systems.^11,12^

The absence of evidence to support the CPAP and HFNO use in patients with COVID-19 led to significant variability both in international guidelines and clinical practice.^9,13^ On this basis, there was an urgent need to determine whether CPAP and HFNO were clinically effective, compared with conventional oxygen therapy, in hospitalized patients with COVID-19 acute respiratory failure.

## METHODS

### Trial design

Recovery-Respiratory Support was a parallel group, open-label, three-arm, adaptive, randomized controlled trial designed to evaluate the clinical effectiveness of CPAP and HFNO, compared with conventional oxygen therapy, in hospitalized patients with acute respiratory failure due to COVID-19. The adaptive design allowed the study to stop early if one or both interventions were more effective than conventional oxygen therapy, with the final analysis adjusted to control the overall alpha value (5%).

The trial protocol was approved by the London-Brighton & Sussex Research Ethics Committee and the Health Research Authority, sponsored by the University of Warwick, co-ordinated by Warwick Clinical Trials Unit, and funded and prioritized as an urgent public health COVID-19 study by the National Institute for Health Research. An independent Trial Steering Committee (TSC) and Data Monitoring Committee (DMC) provided trial oversight. The study was conducted in accordance with Good Clinical Practice guidelines, local regulations, and the ethical principles described in the Declaration of Helsinki.

Consent from patients or agreement from their surrogates was obtained in keeping with regional regulations.

The trial was prospectively registered (ISRCTN16912075) and its design has been published previously.^14^ The trial protocol and statistical analysis plan are available at https://warwick.ac.uk/fac/sci/med/research/ctu/trials/recovery-rs/

### Patients

Adult (≥18-years) hospitalized patients with known or suspected COVID-19 were eligible if they had acute respiratory failure, defined as peripheral oxygen saturations (SpO2) of 94% or below despite receiving a fraction of inspired oxygen (FiO2) of at least 0.4, and were deemed suitable for tracheal intubation if treatment escalation was required. We excluded patients with an immediate (<1-hour) need for invasive ventilation, known pregnancy, or planned withdrawal of treatment. A contraindication to an intervention, based on the judgement of the treating clinician, precluded randomization to that trial arm.

### Randomization

Eligible participants were randomized using an internet-based system with allocation concealment. We anticipated that either CPAP or HFNO might be unavailable at sites on a temporary or permanent basis. As such, the randomization system allowed the treating clinician to randomize between CPAP, HFNO, and conventional oxygen therapy (on a 1:1:1 basis), or between a single intervention (CPAP/HFNO) and conventional oxygen therapy (on a 1:1 basis). Sites could not randomize between CPAP and HFNO only. Randomization was stratified by site, sex, and age, and the allocation was generated by a minimization algorithm.

Participants randomized to CPAP or HFNO started treatment as soon as possible. Breaks from treatment were permitted for comfort. Participants randomized to conventional oxygen therapy continued to receive oxygen via a face mask or nasal cannulae. In all participants, local policies, and clinical discretion informed decisions regarding choice of device, set-up, titration, and discontinuation of treatment. Tracheal intubation was performed when clinically indicated, based on the judgement of the treating clinician. We defined crossover as a participant receiving CPAP or HFNO for more than 6 hours, when not randomized to that intervention, unless it was for the purpose of clinical stabilization, as a bridge to tracheal intubation, or for palliative care.

### Data collection and procedures

At enrolment, we collected information on demographics, co-morbid state, and physiological observations. Participants were followed up throughout their hospital stay to record intervention use, crossover, safety events, and outcomes. We undertook data linkage with national datasets to support collection of demographic information and outcomes. Due to the nature of the trial interventions and context, we were unable to blind patients, treating clinicians, or outcome assessors.

### Outcome measures

The primary outcome was a composite outcome of tracheal intubation or mortality within 30-days of randomization. Tracheal intubation, as an outcome, reflects the need for invasive mechanical ventilation, which is typically delivered in high-resource intensive care units. Secondary outcomes included the incidence of tracheal intubation and mortality at 30 days, time to tracheal intubation, duration of invasive mechanical ventilation, time to death, mortality (critical care, hospital), incidence of intensive care unit admission, and length of stay (critical care, hospital).

### Sample size

Data from Baud et al ^15^ informed the conventional oxygen therapy rate. Assuming a conservative incidence of 15% for the composite outcome of intubation or mortality (with a two-sided 5% significance level and 90% power), a total of 3,000 participants (1,000 per arm across 3 arms) were required. This equated to detecting a reduction of 5% or an odds ratio of 0.625. We further inflated this sample size to 4,002, due to the uncertainties underpinning the sample size assumptions.

### Interim Analyses

Effectiveness monitoring of each pairwise comparison with conventional oxygen therapy was based on an alpha spending function approach with one-sided pairwise type I error rate of 0.025. This allowed the trial to adapt or stop early if one or both interventions were more effective than conventional oxygen therapy. Any decision to stop the trial or drop an arm due to futility or safety was left to the DMC. The sample size calculation assumed 11 interim analyses (and the final one) would be conducted. Only 3 formal interim analyses were conducted at 36, 160 and 387 patients.

### Statistical analysis

The primary and secondary analyses were performed for the intention-to-treat (ITT) population. Outcome data were compared between each intervention arm and conventional oxygen therapy, according to device availability at randomisation. Continuous data were summarized using number of participants, mean, standard deviation (SD), median, and interquartile range (IQR). Categorical data were summarized with frequency count, percentage and missing. Odds ratio (95% confidence interval (CI)) were reported for categorical outcomes using logistic regression models and mean difference (95% CI) were reported for continuous outcomes using linear regression models. For time to event analysis, hazard ratio (95% CI) were reported. The number needed to treat (NNT) was obtained for the primary outcome. Where the 95% CI reflected NNT as infinite, number needed to harm was reported. In the adjusted analyses, covariates age, sex, morbid obesity, ethnicity, FiO2, respiratory rate and treatment phases (baseline variables) were used, with site included as a random effect. Treatment phases were defined as before July 2020, July 2020 to January 2021, after January 2021, based on the introduction of Dexamethasone and Tocilizumab as standard care in June 2020 and January 2021, respectively.^16-18^ Due to the non-availability of NHS Digital data, social deprivation could not be included in the adjusted analyses. The final P value for the primary analysis was corrected for the interim analyses performed.^19^ Thus, P <0.05 was considered as statistically significant for the primary, secondary, and sub-group analyses. Analyses were conducted using SAS, Stata, or R.

## RESULTS

Trial recruitment was stopped to coincide with the end of the funded recruitment period, a rapid decline in UK COVID-19 case numbers, and the need to share accumulated data to inform international treatment of COVID-19 patients. This decision was accepted by the TTSC and agreed by the study sponsor. The trial stopped recruitment on 3^rd^ May 2021.

### Patients

Over the trial period, recruitment closely tracked the number of hospitalized COVID-19 patients in the UK (supplementary information). Recruitment opened at 75 UK hospitals. Between April 2020 and May 2021, there were 1277 randomizations across 48 UK hospitals. Five cases underwent double randomisation, leaving 1272 participants (380 CPAP; 417 HFNO; 475 conventional oxygen therapy) (Figure 1). Eight participants withdrew and five patients were lost to follow-up. Primary outcome data were available for 99.0 % (1259/1272) of participants. We included 733 participants (377 CPAP; 356 conventional oxygen therapy) in the comparison of CPAP with conventional oxygen therapy, and 782 participants (414 HFNO; 368 conventional oxygen therapy) in the comparison of HFNO with conventional oxygen therapy (supplementary information).

**Figure 1:**
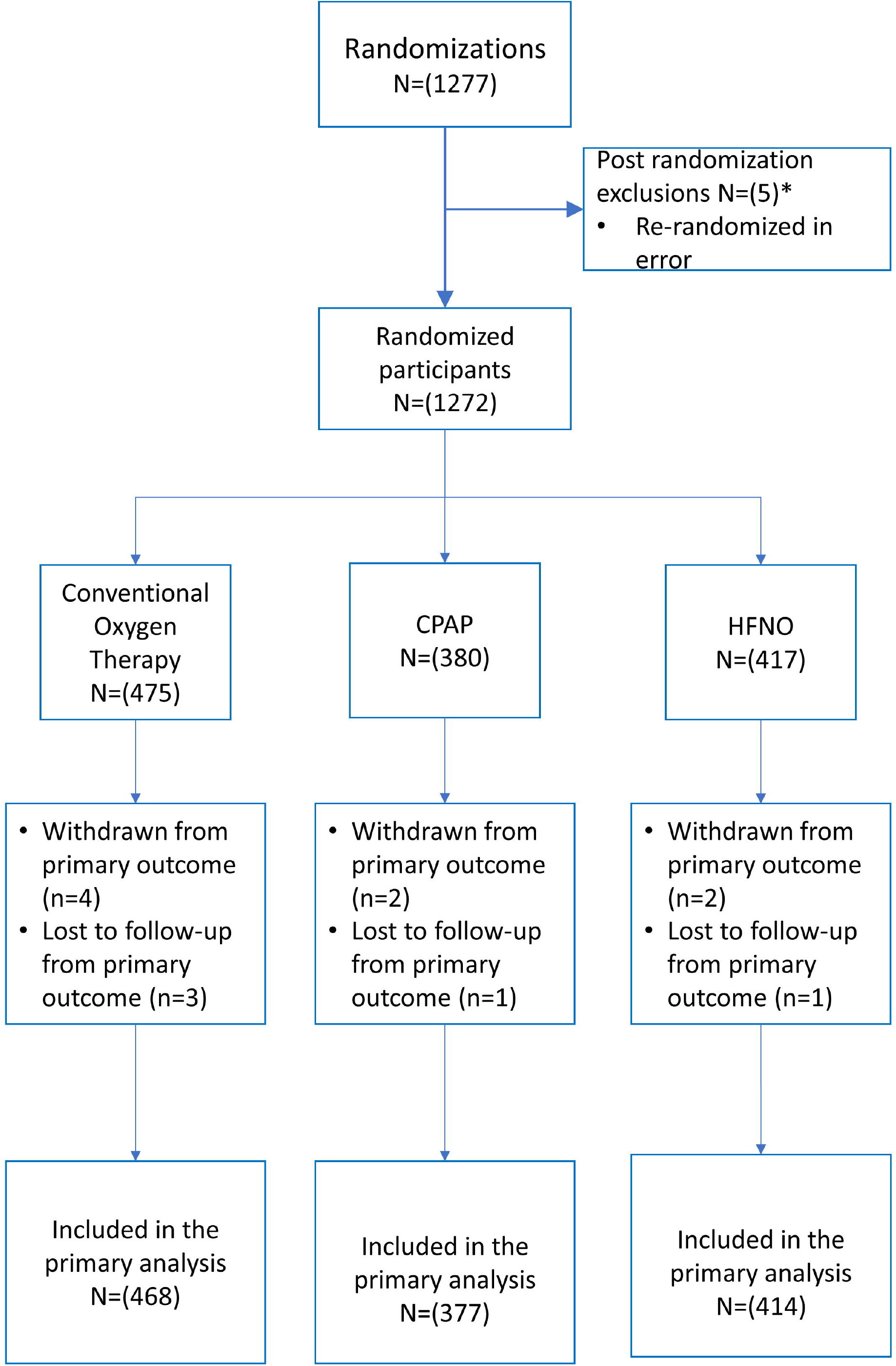
Enrolment and outcomes. Key- CPAP- Continuous Positive Airway Pressure; HFNO-High-flow nasal oxygen * Of the 1272 patients randomized, 5 of these were re-randomized in error and excluded from the summaries and analysis

Participant characteristics were similar at baseline (table 1; supplementary information). The mean age was 57.4 (95% CI, 56.7 to 58.1) years, 66.4% were male, and 65.3% of white ethnicity. Median time from first COVID-19 symptoms to randomization was 9 days (IQR, 7.0 to 12.0). Baseline mean SpO2 and FiO2 were 92.8% (95% CI, 92.6 to 93.0) and 0.61 (95% CI, 0.60 to 0.62) respectively.

**Table 1:** Characteristics of participants at baseline.

| Characteristic | Conventional<br>Oxygen Therapy | CPAP | HFNO | P value |
| --- | --- | --- | --- | --- |
| Mean age ± SD - years | 57.6 ± 12.7 | 56.7 ± 12.5 | 57.6 ± 13.0 | 0.515* |
| Sex – no. (%) |  |  |  | 0.589<sup>#</sup> |
| Male | 312 (65.7) | 260 (68.4) | 272 (65.2) |  |
| Female | 163 (34.3) | 120 (31.6) | 145 (34.8) |  |
| Ethnicity – no. (%) |  |  |  | 0.960<sup>#</sup> |
| White | 312 (65.7) | 243 (64.0) | 275 (66.0) |  |
| Black | 19 (4.0) | 16 (4.2) | 14 (3.4) |  |
| Asian | 90 (19.0) | 73 (19.2) | 77 (18.5) |  |
| Mixed | 6 (1.3) | 3 (0.8) | 4 (1.0) |  |
| Other | 9 (1.9) | 11 (2.9) | 12 (2.9) |  |
| Unknown | 35 (7.4) | 33 (8.7) | 34 (8.2) |  |
| Time from symptom<br>onset to hospital<br>admission (days) |  |  |  |  |
| No. of patients | 466 | 376 | 404 |  |
| Median (IQR) – days | 7.0 (5.0-10.0) | 7.0 (5.5-10.0) | 8.0 (5.0-10.0) | 0.610<sup>¶</sup> |
| Time from symptom<br>onset to randomization<br>(days) |  |  |  |  |
| No. of patients | 470 | 375 | 412 |  |
| Median (IQR) – days | 9.0 (6.0-12.0) | 9.0 (7.0-12.0) | 9.0 (7.0-12.0) | 0.792<sup>¶</sup> |
| COVID-19 status – no. (%) |  |  |  | 0.972<sup>#</sup> |
| Confirmed | 409 (86.1) | 326 (85.8) | 355 (85.1) |  |
| Suspected | 64 (13.5) | 53 (14.0) | 61 (14.6) |  |
| Co-morbidities– no. (%) |  |  |  |  |
| None | 188 (39.6) | 148 (39.0) | 140 (33.6) | 0.385<sup>#</sup> |
| ESRF requiring RRT | 5 (1.1) | 2 (0.5) | 6 (1.4) | 0.783<sup>#</sup> |
| Congestive cardiac<br>failure | 5 (1.1) | 2 (0.5) | 4 (1.0) | 0.932<sup>#</sup> |
| Chronic lung disease | 66 (13.9) | 65 (17.1) | 52 (12.5) | 0.445<sup>#</sup> |
| Coronary heart disease | 44 (9.3) | 34 (9.0) | 26 (6.2) | 0.515<sup>#</sup> |
| Dementia | 3 (0.6) | 4 (1.1) | 1 (0.2) | 0.699<sup>#</sup> |
| Diabetes requiring<br>medication | 91 (19.2) | 86 (22.6) | 98 (23.5) | 0.581<sup>#</sup> |
| Hypertension | 153 (32.2) | 131 (34.5) | 164 (39.3) | 0.228<sup>#</sup> |
| Uncontrolled or active<br>malignancy | 7 (1.5) | 7 (1.8) | 10 (2.4) | 0.890<sup>#</sup> |
| Morbid obesity (BMI<br>>35) | 75 (15.8) | 62 (16.3) | 81 (19.4) | 0.623<sup>#</sup> |
| Clinical Frailty Scale (pre-<br>admission) – no. (%) |  |  |  | 0.079<sup>#</sup> |
| CFS1 - Very Fit | 62 (13.1) | 72 (19.0) | 70 (16.8) |  |
| CFS2 - Well | 237 (49.9) | 192 (50.5) | 196 (47.0) |  |
| CFS3 - Managing Well | 131 (27.6) | 87 (22.9) | 109 (26.1) |  |
| CFS4 - Vulnerable | 30 (6.3) | 12 (3.2) | 27 (6.5) |  |
| CFS5 - Mildly Frail | 6 (1.3) | 4 (1.1) | 6 (1.4) |  |
| CFS6 - Moderately Frail | 3 (0.6) | 3 (0.8) | 0 (0.0) |  |
| CFS7 - Severely Frail | 0 (0.0) | 0 (0.0) | 2 (0.5) |  |
| CFS8 - Very Severely Frail | 0 (0.0) | 0(0.0) | 0 (0.0) |  |
| CFS9 - Terminally Ill | 0 (0.0) | 0 (0.0) | 0 (0.0) |  |
| Respiratory rate |  |  |  |  |
| No. of patients | 472 | 377 | 414 |  |
| Mean ± SD – (breaths per minute) | 25.0 ± 6.8 | 26.4 ± 7.5 | 25.4 ± 7.0 | 0.017 * |
| FiO<sub>2</sub> |  |  |  |  |
| No. of patients | 458 | 362 | 404 |  |
| Mean ± SD | 0.61 ± 0.24 | 0.62 ± 0.24 | 0.60 ± 0.24 | 0.633 * |
| SpO<sub>2</sub> |  |  |  |  |
| No. of patients | 471 | 377 | 409 |  |
| Mean ± SD – (%) | 93.1 ± 3.8 | 92.9 ± 3.7 | 92.5 ± 4.3 | 0.076 * |
| SpO<sub>2</sub> to FiO<sub>2</sub> ratio |  |  |  |  |
| No. of patients | 457 | 361 | 400 |  |
| Mean ± SD – (%) | 186.4 ± 99.1 | 182.8 ± 94.7 | 186.3 ± 97.5 | 0.841 * |
| PaO<sub>2</sub> |  |  |  |  |
| No. of patients | 315 | 238 | 284 |  |
| Test not available | 140 | 122 | 108 |  |
| Mean ± SD (mmHg) | 73.3 ± 24.2 | 71.0 ± 17.8 | 70.0 ± 20.1 | 0.150 * |
| PaO<sub>2</sub> to FiO<sub>2</sub> ratio |  |  |  |  |
| No. of patients | 305 | 228 | 281 |  |
| Test not available | 140 | 122 | 108 |  |
| Mean ± SD | 134.9 ± 82.8 | 131.8 ± 67.8 | 138.5 ± 87.6 | 0.643 * |
| Awake prone positioning – no. (%) |  |  |  | 0.014 # |
| Yes | 252 (53.1) | 207 (54.5) | 243 (58.3) |  |
| No | 122 (25.7) | 120 (31.6) | 98 (23.5) |  |
| Unknown | 90 (19.0) | 51 (13.4) | 72 (17.3) |  |
| CPAP set-up PEEP |  |  |  |  |
| No. of patients | - | 312 | - |  |
| Mean ± SD (cmH<sub>2</sub>O) | - | 9.5 (8.4) | - | |
| HFNO set-up flow |  |  |  |  |
| No. of patients | - | - | 336 |  |
| Mean ± SD (L/min) | - | - | 50.8 (12.6) | |

Key- BMI- body mass index; CPAP- Continuous Positive Airway Pressure; ESRF- end-stage renal failure; FiO<sub>2</sub>- fraction of inspired oxygen; HFNO- High-flow nasal oxygen; PaO<sub>2</sub>-Partial pressure of oxygen; PEEP- Positive End Expiratory Pressure; RRT- Renal replacement therapy; SpO- Peripheral oxygen saturation;
\* T-test used to calculate p-value; # Chi-squared statistics used to calculate the p-value; ¶ Kruskal Wallis test was used to calculate the p-value

The allocation intervention was received by 348/380 (91.6%), 384/417 (92.1%), and 467/475 (98.3%) participants in the CPAP, HFNO, and conventional oxygen arms, respectively (supplementary information). Crossover occurred in 58/380 (15.3%) of participants in the CPAP arm, 48/417 (11.5%) in the HFNO arm, and 112/475 (23.6%) in the conventional oxygen therapy arm.

### Primary outcome

For the comparison of CPAP and conventional oxygen therapy, the primary outcome occurred in 137/377 (36.3%) participants in the CPAP group and 158/356 (44.4%) participants in the conventional oxygen therapy group (unadjusted odds ratio 0.72, 95% CI 0.53 to 0.96, P=0.03). For the comparison of HFNO and conventional oxygen therapy, the primary outcome occurred in 184/414 (44.4%) participants in the HFNO group and 166/368 (45.1%) participants in the conventional oxygen therapy group (unadjusted odds ratio 0.97; 95% CI 0.73 to 1.29, P=0.85). Findings were consistent across both unadjusted and adjusted analyses (Table 2).

**Table 2:** Primary and Secondary outcomes.

| Outcome | Pairwise Treatment Comparisons |  |  |  | Odds Ratio†/Hazard Odds‡/Mean Difference§ (95% CI) |  |  |  |
| --- | --- | --- | --- | --- | --- | --- | --- | --- |
|  | CPAP versus Conventional Oxygen Therapy¶ |  | HFNO versus Conventional Oxygen Therapy\|\| |  | CPAP versus Conventional Oxygen Therapy¶ |  | HFNO versus Conventional Oxygen Therapy\|\| |  |
|  | CPAP | Conventional Oxygen Therapy | HFNO | Conventional Oxygen Therapy | Unadjusted | Adjusted | Unadjusted | Adjusted |
| Tracheal Intubation or mortality within 30 days – no./total (%)† | 137/377 (36.3) | 158/356 (44.4) | 184/414 (44.4) | 166/368 (45.1) | 0.72 (0.53- 0.96) | 0.67 (0.48- 0.94) | 0.97 (0.73 -1.29) | 0.95 (0.69- 1.30) |
| Intubation within 30 days – no./total (%)† | 126/377 (33.4) | 147/356 (41.3) | 170/414 (41.1) | 153/368 (41.6) | 0.71 (0.53- 0.96) | 0.66 (0.47- 0.93) | 0.98 (0.74- 1.30) | 0.96 (0.70 - 1.31) |
| Mortality at 30 days – no./total (%)† | 63/378 (16.7) | 69/359 (19.2) | 78/415 (18.8) | 74/370 (20.0) | 0.84 (0.58-1.23) | 0.91 (0.59 -1.39) | 0.93 (0.65 -1.32) | 0.96 (0.64 - 1.45) |
| **Secondary outcomes #** |  |  |  |  |  |  |  |  |
| Tracheal Intubation rate in the study period – no./total (%)†, * | 126/377 (33.4) | 147/356 (41.3) | 169/414 (40.8) | 154/368 (41.8) | 0.71 (0.53- 0.96) | 0.66 (0.47- 0.93) | 0.96 (0.72- 1.28) | 0.93 (0.68- 1.28) |
| Admission to critical care – no./total (%)† | 205/379 (54.1) | 219/356 (61.5) | 253/416 (60.8) | 214/368 (58.2) | 0.74 (0.55- 0.99) | 0.69 (0.49- 0.96) | 1.12 (0.84- 1.49) | 1.06 (0.76-1.47) |
| Median duration of invasive ventilation (IQR) - days |  |  |  |  |  |  |  |  |
| All randomized patients‡ | 0.0 (0.0 - 7.0) | 0.0 (0.0 - 8.4) | 0.0 (0.0 - 10.0) | 0.0 (0.0 - 8.8) | NA | NA | NA | NA |
| Intubated patients ‡ | 14.0 (8.0 - 24.0) | 10.0 (5.0 - 21.0) | 14.0 (6.0 - 25.0) | 11.0 (5.0 - 21.0) | 0.76 (0.56- 1.04) | 0.79 (0.57-1.08) | 0.93 (0.72 – 1.21) | 1.00 (0.75 - 1.33) |
| Median time to intubation (IQR) – days ‡ | 2.2 (1.0 – 4.6) | 1.0 (0.0-4.0) | 1.0 (0.0-4.0) | 1.0 (0.0-3.0) | 0.74 (0.58 -0.94) | 0.67 (0.52 - 0.86) | 0.96 (0.77 - 1.20) | 0.91 (0.72 -1.14) |
| Median time to death (IQR) – days ‡ | 17.0 (11.0- 26.0) | 17.0 (11.0-24.0) | 16.5 (9.0-22.5) | 17.0 (11.0-24.0) | 0.86 (0.61-1.21) | 0.93 (0.65-1.33) | 0.94 (0.68- 1.29) | 0.94 (0.67 - 1.31) |
| Mortality in critical care † | 62/204 (30.4) | 65/219 (29.7) | 72/251 (28.7) | 64/214 (29.9) | 1.03 (0.68-1.57) | 1.13 (0.72- 1.80) | 0.94 (0.63-1.41) | 1.00 (0.63 -1.56) |
| Mortality in hospital † | 72/364 (19.8) | 78/346 (22.5) | 88/404 (21.8) | 80/359 (22.3) | 0.85 (0.59 - 1.22) | 0.92 (0.61 -1.37) | 0.97 (0.69 -1.37) | 1.02 (0.69 -1.50) |
| Mean length of stay in critical care (SD) – days § | 9.5 (15.6) | 9.6 (13.6) | 10.5 (15.6) | 9.5 (14.1) | -0.08 (-2.23, 2.07) | -0.33 (-2.44, 1.78) | 1.01 (-1.11, 3.14) | 0.69 (-1.37, 2.75) |
| Mean length of stay in hospital<br>(SD) – days § | 16.4 (17.5) | 17.3 (18.1) | 18.3 (20.0) | 17.1 (18.0) | -0.96 (-3.59, 1.67) | -0.97 (-3.65,<br>1.71) | 1.25 (-1.46, 3.97) | 0.70 (-1.93, 3.34) |

Key- CPAP- Continuous Positive Airway Pressure; HFNO- High-flow nasal oxygen
The % are based on excluding missing data (i.e. withdrawals and no data provided).

The number needed to treat for CPAP was 12 (95% CI, 7 to 105) and for HFNO was 151 (95% CI, number needed to treat 13 to number needed to harm 16).

### Secondary outcomes

Secondary outcomes are presented in Table 2. The decrease in the primary outcome in the CPAP group was driven by a decrease in the incidence of tracheal intubation, with no statistically significant difference in rate of 30-day mortality (Table 2). Neither CPAP nor HFNO, compared with conventional oxygen therapy, reduced mortality at any time-point. In the CPAP group, fewer participants required admission to critical care and, in those that required tracheal intubation, time to tracheal intubation was longer (Figure 2). There was no significant difference for any other outcome in the comparison of CPAP and conventional oxygen therapy or for any outcome in the comparison of HFNO and conventional oxygen therapy.

**FIGURE 2:**
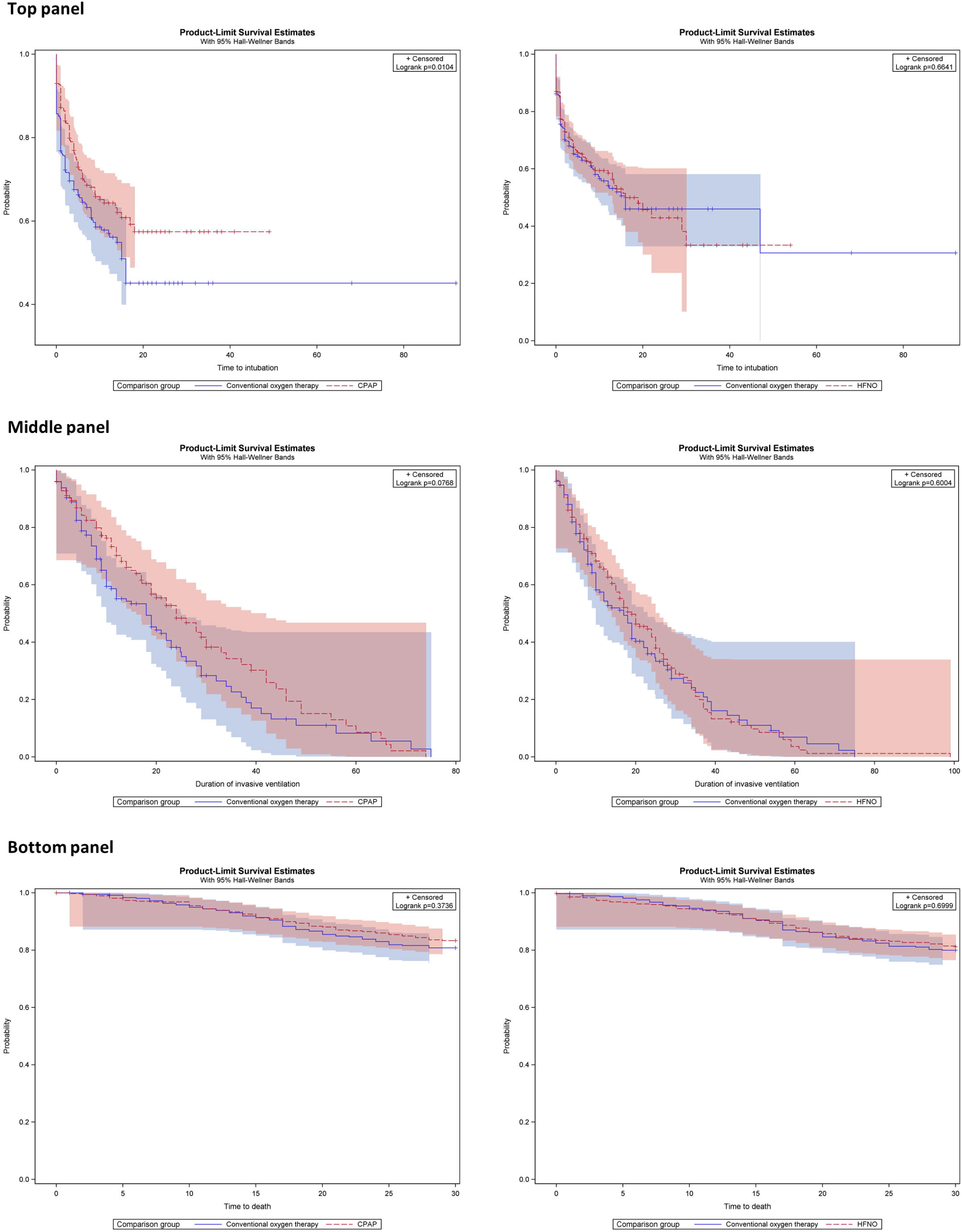
Kaplan Meier curves by treatment arm: top pane-time to tracheal intubation (all participants); middle panel-duration of invasive ventilation (intubated participants only); bottom panel-survival (all participants). For all panels, left side shows CPAP vs conventional oxygen therapy; right side shows HFNO vs conventional oxygen therapy Note: Time to intubation: censored patients include 1) patients died before hospital discharge; 2) patients discharged alive from hospital without intubation; 3) patients withdrew completely before intubation was given. Duration of invasive ventilation: censored patients include: 1) patients died before coming off invasive ventilation alive; 2) patients withdrew completely before the end of invasive ventilation; 3) patients stayed on invasive ventilation beyond the follow-up period (20^th^ June, 2021); 4) patient died in hospital or discharged alive without intubation. Time to death: censored patients include: 1) patient withdrew completely before 30 days from randomisation; 2) patient survived at 30 days

### Safety

Safety events, as summarized in the supplementary information, were most commonly reported in the CPAP group (CPAP 130/380 (34.2%); HFNO 86/417 (20.6%); conventional oxygen therapy 66/475 (13.9%), p<0.001). The most commonly reported adverse event was hemodynamic instability, occurring in 108 (8.5%) participants. Across all groups, pneumothorax and pneumomediastinum events were reported in 26 (2.0%) and 20 (1.6%) participants respectively. Eight serious adverse events (seven CPAP; one conventional oxygen therapy) were reported. Four were classified as probably or possibly linked to the trial intervention, with all occurring in the CPAP group (surgical emphysema and pneumomediastinum; pneumothorax and pneumomediastinum (two events); and vomiting requiring emergency tracheal intubation).

## DISCUSSION

In this open-label, three-arm, adaptive, randomized controlled trial, we included hospitalized adults with acute respiratory failure due to COVID-19 deemed suitable for tracheal intubation if treatment escalation was required. We found that CPAP, compared with conventional oxygen therapy, was effective in reducing the composite outcome of tracheal intubation or mortality within 30-days. In contrast, HFNO provided no benefit compared with conventional oxygen therapy. This decrease in the incidence of the primary outcome with CPAP was attributable to a decrease in the need for tracheal intubation. Neither HFNO nor CPAP reduced mortality, compared with conventional oxygen therapy.

We designed a pragmatic trial that was deliverable in the context of a pandemic and which tested interventions that precluded blinding of either the participant or treating clinician. The decision to commence invasive mechanical ventilation was not standardised.^13^ As such, the lower rate of tracheal intubation seen with CPAP may have been driven by a greater willingness amongst clinicians and patients to delay intubation. Our finding that time to tracheal intubation was longer in the CPAP group may support this proposition. However, this same effect was not observed with HFNO which should have been susceptible to the same risk of performance bias. Protocolising intervention delivery and escalation to tracheal intubation, as in the HENIVOT trial, may have mitigated this issue but at the expense of pragmatism, generalizability, and deliverability across a large number of hospitals during a pandemic.^20^

A recent systematic review and meta-analysis of 25 randomized controlled trials (3804 patients) summarized evidence on the clinical effectiveness of non-invasive ventilation (with and without pressure support) and HFNO, compared with conventional oxygen therapy, in acute respiratory failure.^5^ Key findings were that non-invasive ventilation reduced the incidence of both tracheal intubation and mortality, whilst HFNO reduced only the incidence of tracheal intubation. We found that CPAP reduced tracheal intubation, but not mortality, although our trial was not specifically powered to detect differences in mortality. We found that HFNO did not reduce the need for tracheal intubation. One explanation for these discordant findings is differences in pathophysiology between COVID pneumonitis and other causes of acute respiratory failure^5,21^ Furthermore, in our trial, some hospitals modified care pathways to deliver CPAP and HFNO outside of a critical care unit, which may have influenced the clinical effectiveness of both interventions.

Two randomised controlled trials of non-invasive respiratory strategies in COVID-19 have previously reported.^20,22^ One trial of 22 patients that compared HFNO with conventional oxygen therapy reported that HFNO improved PaO_2_/FiO_2_ ratio and reduced ICU length of stay.^22^ These data should be interpreted with caution due to the small sample size and high risk of bias. In contrast to our study, the HENIVOT trial directly compared non-invasive ventilation with pressure support via a helmet and HFNO in 110 COVID-19 patients.^20^ In the HENIVOT trial, no difference was observed in the primary outcome of days free of respiratory support, although fewer patients in the non-invasive ventilation arm required tracheal intubation (odds ratio 0.41, 95% CI 0.18-0.89). However, the trial’s highly protocolised approach to the set-up and weaning of trial interventions and the decision to perform tracheal intubation potentially limits its generalizability.

Our trial has several limitations. Firstly, we did not achieve our planned sample size with the decision to stop recruitment driven by practical reasons linked to reducing numbers of COVID-19 in the UK, and an ethical obligation to share accumulated data with the international clinical community. Secondly, we observed crossover between allocated treatment arms, principally from the conventional oxygen therapy arm to one or both of the interventions. This is a common challenge in trials of non-invasive respiratory strategies, and reduces the observed effect size of a clinically effective treatment.^23,24^ Thirdly, we determined that it would be impractical to collect screening data, meaning we are unable to describe the number of non-randomized patients and reasons for non-randomization. Finally, the trial was rapidly set-up early in the pandemic, prior to the development of a core outcome set for COVID-19 trials.^25^ Whilst our outcome list aligns closely to most of the core outcomes subsequently identified, we did not capture information on patient recovery following hospital discharge.

In conclusion, in this randomized trial of hospitalized adults with acute respiratory failure due to COVID-19, CPAP, compared with conventional oxygen therapy, reduced the composite outcome of tracheal intubation or death within 30 days of randomisation in hospitalized adults with acute respiratory failure due to COVID-19. There was no effect observed, compared with conventional oxygen therapy, with the use of HFNO.

## Supporting information

electronic supplement

CONSORT checklist

## Data Availability

Requests for data sharing will be reviewed on an individual basis by the chief investigators. The study will comply with the good practice principles for sharing individual participant data from publicly funded clinical trials and data sharing will be undertaken in accordance with the required regulatory requirements.

https://warwick.ac.uk/fac/sci/med/research/ctu/trials/recovery-rs/

## Data Availability

All the data are included in the manuscript and available upon requests.

## Declaration

This study is funded by the National Institute for Health Research (NIHR) [COVID-19-RSC]. The views expressed are those of the author(s) and not necessarily those of the NIHR or the Department of Health and Social Care. The funder had no role in the trial design, in the collection or analysis of the data, or in the writing of the manuscript.

Professor Perkins is supported as an NIHR senior investigator, through the NIHR West Midlands Applied Research Collaboration. Professor McAuley is programme director for the NIHR Efficacy and Mechanism Evaluation programme. Professor Perkins and Dr Connolly, are directors of research for the Intensive Care Society. Professor McAuley was, until recently (term ended June 2021), a director of research for the Intensive Care Society Professor Dark is NIHR CRN National Specialty Cluster Lead and is also supported by the Manchester NIHR Biomedical Research Centre.

Mrs Devrell reports personal fees from the NIHR for patient and public involvement work related to the study. Outside of the submitted work, the following conflicts of interest were declared. Dr Connolly reports grant funding from the NIHR and personal fees from Fisher and Paykel. Dr Dave reports personal fees from Chesei. Professor De Soyza reports grant support, speaker’s fees, advisory board fees and conference attendance support from AstraZeneca, Bayer, Chiesi, Gilead, GlaxoSmithKline, Pfizer, Forest labs, Novartis, Insmed, and Zambon. Professor Hart reports grant funding from the NIHR, UK Research and Innovation, with unrestricted grants and equipment from Philips-Respironics, Fisher and Paykel, and Resmed; financial support from Philips for development of the MYOTRACE technology that has patent approved in Europe and US; personal fees for lecturing from Philips-Respironics, Philips, Resmed, and Fisher and Paykel; and institutional funding for his role on the Philips Global Medical Advisory Board. Dr Messer reports personal fees from Fisher and Paykel. Dr Parekh reports grant funding from the NIHR and Medical Research Council UK Research and Innovation. Professor Steiner reports personal fees from GlaxoSmithKline. Professor McAuley reports personal fees from consultancy for GlaxoSmithKline, Boehringer Ingelheim, Bayer, Novartis, SOBI and Eli Lilly, and from sitting on DMECs for trials undertaken by Vir Biotechnology and Faron Pharmaceuticals. Professor McAuley also reports grant funding to his institution from several funders (NIHR, Wellcome Trust, Innovate UK, Medical Research Council, and Northern Ireland Health and Social Research and Development division) for studies in patients with ARDS and COVID-19, and a patent (US8962032) issued to his institution as a treatment for inflammatory disease. The remaining authors report no conflicts of interest.

We are grateful to all the patients and families who supported the trial, together with the doctors, nurses, and allied health professionals across all participating hospitals who supported both trial recruitment and delivery of trial interventions in extremely challenging conditions. We thank the NIHR Clinical Research Network and Northern Ireland Clinical Research Network. We also thank Health Data Research UK, the Office for National Statistics and the Intensive Care National Audit and Research Centre for support with data linkage. Finally, we are indebted to the members of both the trial steering committee and the data monitoring committee, namely: Professor Kathy Rowan, Professor Duncan Young, Professor Marion Campbell, Susie Hennings, Professor John Laffey, Professor Martin Landray, Gillian McCarmack, Gary Overton, Dr Marion Thompson, Professor Taylor Thompson, and Professor Tim Walsh.

