## Supplementary material for "An adaptive randomized controlled trial of non-invasive respiratory strategies in acute respiratory failure patients with COVID-19": electronic supplement

Contents:


| | |
|---|---|
| Recovery- RS collaborators | Page 2 |
| FIGURE S1: RECOVERY RS trial recruitment and UK hospitalized COVID-19 patients | Page 9 |
| Table S1: Summary of randomizations and data for pairwise comparisons | Page 10 |
| Table S2: Summary of trial crossover by treatment arm | Page 11 |
| Table S3: Additional participant baseline characteristics | Page 12 |
| Table S4: Adverse events and serious adverse events by treatment arm | Page 13 |

### Recovery- RS collaborators

**Aintree University Hospital, Liverpool University Hospitals NHS Foundation Trust, Liverpool, UK**

Dr Nick Duffy (PI).

**Altnagelvin Area Hospital, Western Health and Social Care Trust, Londonderry, UK**

Dr Martin Kelly (PI), Donal Concannon, Kathryn Ferguson, Declan McClintock.

**Barnet Hospital, Royal Free London NHS Foundation Trust, London, UK**

Dr Rajeev Jha (PI), Vinodh Krishnamurthy, Stephen O'Farrell.

**Belfast City Hospital, Belfast Health and Social Care Trust, Belfast, UK**

Prof Cecilia O'Kane (PI).

**Charing Cross Hospital, Imperial College Healthcare NHS Trust, London, UK**

Dr Clare Ross (PI), Dr Richard Douglas Turner, Serge Miodragovic.

**Colchester General Hospital, East Suffolk and North Essex NHS Foundation Trust, Colchester, UK**

Dr Peter Hawkins (PI).

**Derriford Hospital, University Hospitals Plymouth NHS Trust, Plymouth, UK**

Dr Jessie Welbourne (PI), Colin Wells, Liana Lankester, Dr Samuel David Waddy, Dr Julian Lentaigne.

**Fairfield General Hospital, North care Alliance NHS Group**

Dr Jay Nesbitt (PI), Dr Sarah Clarke, Dr Catherine Houghton, Dr Devaki O'Riordan, Dr Kate Shepherd, Dr Beth Turnpenny, Rosane Joseph.

**Glenfield Hospital, University Hospitals of Leicester NHS Foundation Trust, Leicester, UK**

Professor Michael Steiner (PI), Clare Rossall, Rachel Mundin, Samuele Boschi, Dr Hamish J C McAuley, Dr Richard J Russell, Dr Sarah Diver, Dr Omer Elneima, Dr Wadah Ibrahim, Dr Ahmed Yousuf, Sarah Edwards.

**Good Hope Hospital, University Hospitals Birmingham NHS Foundation Trust, Birmingham, UK**

Dr Mohammad Saim (PI), Bridget Hopkins, Lisa Kelly, Daniel Lenton, Helen Shackleford, Laura Thrasyvoulou, Heather Willis.

**Grange University Hospital, Aneurin Bevan University Health Board, Cwmbran, UK**

Dr Sara Fairbairn (PI).

**Heartlands Hospital, University Hospitals Birmingham NHS Foundation Trust, Birmingham, UK**

Dr Chris Green (PI), Ed Birkhamshaw, Emma Gallagher, Safia Begum, Louise Wood, Mirriam Sangombe, Will Osborne, Pretin Davda, Kate Clay, Salman Khurshid, Ed Justice, Darryl Menezes, Matt Page, Emma Hayton, Martin Dedicoat, Andrew Woodhouse, Neil Jenkins, Neil Crooks, Lucie Linhartova, Amy Chue, Aimee Johnson, Chris Pollard, Mark Bailey, Matt O'Shea, Jamie Scriven, Mimi Hamad, Jennifer Short, Dan Burns, Jessica Sale, Vivien Price, Kiranmai Bhatt, Azra Parveen, Kush Naker, Jo Whitehouse, Neil Patel, Karishma Gokani, Samira Afzal, Pearlene Antoine-Pitterson, Hollie Bancroft, Sundip Dhani, Mary Bellamy, Heather Willis, Bridget Hopkins, Daniel Lenton, Helen Shackleford, Lisa Kelly, Laura Thrasyvoulou, Beth Jones.

**Hull Royal Infirmary, Hull University Teaching Hospitals NHS Trust, Hull, UK**

Dr Michael Crooks (PI), Kayleigh Brindle, Dr Shoaib Faruqi, Rachel Flockton, Dr Emma Pinder, Susannah Thackray-Nocera.

**Intensive Care National Audit and Research Centre**

Keji Dalemo, Dr James Doidge, Dr Julia Edwards

**Ipswich Hospital, East Suffolk and North Essex NHS Foundation Trust, Ipswich, UK**

Dr Jonathon Douse (PI), Stephanie Bell, Bally Purewal, Cathleen Chabo, Carol Buckman, Deborah Beeby, Georgina Gray, Rebecca Francis, Vanessa Rivers, , Dr Matthew Burton, Dr Nicholas Innes, Dr Sandy Ghattas, Dr Rana Rabbani.

**James Paget University Hospital, James Paget University Hospitals NHS Foundation Trust, Great Yarmouth, UK**

Dr Venkat Mahadevan (PI), Dr Venkateswaran Mahadevan, Dr Alastair Green, Prof Ben Burton, Christian Hacon, Elva Wilhelmsen.

**Jersey General Hospital, St Helier, Jersey**

Dr Paul Richard Hughes (PI).

**King's College Hospital, King's College Hospital NHS Foundation Trust, London, UK**

Dr Kai Lee (PI).

**Leighton Hospital, Mid Cheshire Hospitals NHS Foundation Trust, Crewe, UK**

Dr Richard Lowsby (PI), Dr Laurence Baker, Dr Perry Board, Dr Varun Chauhan, Sheron Clarke, Dr Duncan Fullerton, Claire Gabriel, Dr Tom Houston, Dr Diana Lees, Dr Robert Normanton, Katherine Pagett, Dr Sarah Thornley, Dr Harriet Wright.

**Lister Hospital, East and North Hertfordshire NHS Trust, Stevenage, UK**

Dr Alison McMillan (PI).

**Macclesfield District General Hospital, East Cheshire NHS Trust, Macclesfield, UK**

Dr Marta Babores (PI), Dr Xiang Lee, Dr Thapas Nagarajan, Maureen Holland.

**Medway Maritime Hospital, Medway NHS Foundation Trust, Gillingham, UK**

Dr Thomas Sanctuary (PI).

**Musgrove Park Hospital, Somerset NHS Foundation Trust, Taunton, UK**

Dr Richard Innes (PI).

**Norfolk and Norwich University Hospital, Norfolk and Norwich University Hospitals NHS Foundation Trust, Norwich, UK**

Dr Simon Fletcher (PI).

**North Manchester General Hospital, North Care Alliance**

Dr Nita Sehgal (PI), Dr Tracy Duncan.

**Office for National Statistics**

Justine Pooley

**Princess of Wales Hospital (Wales), Cwm Taf Morgannwg University Health Board, Bridgend, UK**

Dr Emma Watkins (PI).

**Princess Royal Hospital, Shrewsbury and Telford Hospital NHS Trust, Teleford, UK**

Dr Harmesh Moudgil (PI), Mandy Carnahan, Denise Donaldson

**Princess Royal University Hospital, King's College Hospital NHS Foundation Trust, Orpington, UK**

Dr Deepak Rao (PI), Dr Chia Ling Tey, Dr Lynette Linkson, Dr Tom Buttle, Dr Jennifer Vidler, Nicola Griffiths.

**Queen Alexandra Hospital, Portsmouth Hospitals University NHS Trust, Portsmouth, UK**

Alexander Hicks (PI), Dr Hitasha Rupani, Afaq Alfridi, Debi Barns, Elena Cowan, Mini David, Alex Darbyshire, Ben Giles, Claire Roberts, Claudia Lameirinhas, Daniel Neville, Ejaz Hossain, Fiona Thompson, Helena Edwards, Jen Naftel, Jonathan Winter, Kate Burrows, Laura Wiffen, Lauren Fox, Lisa Murray, Liz Hawes, Madhu Mamman, Maria Moon, Marie White, Megan Rowley, Nina Szarazova, Sally Gosling, Simon Cooper, Sonia Baryschpolec, Sophie Arndtz, Yasmin H-Davies, Yazeed Abed El Khaleq, Zoe Garner, Siyamini Vythilingam, Yingjja Yang

**Queen Elizabeth Hospital, University Hospitals Birmingham NHS Foundation Trust, Birmingham, UK**

Dr Dhruv Parekh (PI), Dr Shyam Madathil, Dr Jaimin Patel, Colin Bergin, Michelle Bates, Christopher McGhee, Daniella Lynch, Khushpreet Bhandal, Kyriaki Tsakiridou, Amy Bamford, Lauren Cooper, Dr Tony Whitehouse, Dr Tonny Veenith, Elliot Forster, Steph Lane, Nick Adams, Sonia MacDonald, Sana Manan, Dr Sebastian Lugg, Dr Peer Ameen Shah, Dr Emily McKemey, Dr Louise Crowley, Dr Gulfam Mussawar, Dr Atena Gogokhia, Dr Simon Gompertz, Dr Catherine Snelson, Dr Tessa Oelofse, Dr Jeremy Wilson, Dr Mansoor Bangash, Dr Syed Sa Huq, Dr Farrukh Rauf, Dr Davinder Dosanjh, Natasha Salmon, Joyce Tengende, Kay Filby Senior, Prof Brendan Cooper, Dr Benjamin Sutton, Dr Ian Woolhouse, Dr Anjali Crawshaw, Dr Richard Thompson, Dr Patricia Glynn, Dr Jon Naylor, Dr Joseph Alderman, Dr Minesh Chotalia, Dr Martin Le Breuilly, Dr Nicholas Talbot, Dr Gregory Packer.

**Queen Elizabeth University Hospital (Glasgow), Greater Glasgow Health Board, Glasgow, UK**

Dr Chris Carlin (PI).

**Queen's Medical Centre, Nottingham University Hospitals NHS Trust, Nottingham, UK**

Dr Dan Harvey (PI)

**Royal Gwent Hospital, Aneurin Bevan University Health Board, Newport, UK**

Dr Sara Fairbairn (PI).

**Royal Infirmary of Edinburgh, NHS Lothian, Edinburgh, UK.**

Prof. Alasdair Gray.

**Royal Liverpool University Hospital, Liverpool University Hospitals NHS Foundation Trust, Liverpool, UK**

Dr Manish Gautam (PI), Prof Ingeborg Welters (Co-PI), Dr David Oliver Hamilton, Dr Hassan Burhan, Karl Hunter, Dr. Brian Johnston, Maria Lopez, Catherine Lowe, Dr. Suleman Mulla, Jaime Fernandez Roman, David Shaw, Dr. Alicia Waite, Victoria Waugh, Karen Williams.

**Royal Brompton Hospital, Royal Brompton and Harefield NHS Foundation Trust, UK**

Professor Anita K Simonds (PI).

**The Royal Marsden Hospital (London) and The Royal Marsden Hospital (Surrey), The Royal Marsden NHS Foundation Trust, London, UK**

Dr Kate C Tatham (PI), Ethel Black, Shaman Jhanji.

**Royal Oldham Hospital, North Care Alliance NHS Group, Oldham, UK**

Dr Georges Ng Man Kwong (PI).

**Royal Victoria Infirmary, Newcastle Hospitals NHS Foundation Trust, Newcastle upon Tyne, UK**

Dr. Ben Messer (PI), Prof Anthony De-Soyza, Paul McAlinden, Sophie D West.

**Russells Hall Hospital, The Dudley Group NHS Foundation Trust, Dudley, UK**

Dr. Vikram Anumakonda (PI).

**Salford Royal Hospital, Northern Care Alliance NHS Group< Manchester, UK**

Professor Paul Dark, Liam McMorrow, Tracy Marsden, Nicola Proudfoot, Bethan Charles, Jessica Pendlebury, Bethan Blackledge, Alice Harvey, Karen Knowles, Reece Doonan, Stephanie Lee, Jane Perez, Melanie Slaughter, Melanie Taylor, Victoria Thomas, Dr Emma Hardy, Prof Nawar Bakerly, Laura Catlow, Dr Nasir Majeed, Bethan Charles, Prof Dan Horner.

**Scunthorpe General Hospital, North Lincolnshire and Goole NHS Foundation Trust, UK**

Dr Liaquat Ali (PI), Dorothy Hutchinson.

**South Tyneside District Hospital, South Tyneside and Sunderland NHS Foundation Trust, South Shields, UK**

Dr Liz Fuller (PI).

**Southmead Hospital, North Bristol NHS Trust, Bristol, UK**

Dr James Dodd (PI), Dr Rahul Bhatnagar, Dr Amelia Clive, Dr Huzaifa Adamali, Dr Anna Bibby, Dr Daniel Higbee, Dr Hugh Welch, Emma Gendall, Louise Staddon, Anna Morley, Sam Clarke, Kerry Smith, Emily Perry, Naomi Rippon, Louise Jennings, Louise Solomon, Karen Alloway, Hannah Lee, Victoria Sandrey, Kirstie Bradburn, Alice Milne, Elizabeth Goff, Rachel Williams.

**St George's Hospital, St George's University Hospitals NHS Foundation Trust, London, UK**

Dr Mohammed Ahmed (PI).

**St Mary's Hospital, Imperial College Healthcare NHS Trust, London, UK**

Dr Clare Ross (PI), Dr Susannah Bloch, Serge Miodragovic.

**Stepping Hill Hospital, Stockport NHS Foundation Trust, Stockport, UK**

Dr Ahmed Zaki (PI).

**Sunderland Royal Hospital, South Tyneside and Sunderland NHS Foundation Trust, Sunderland, UK**

Dr Ali Roy (PI).

**Southampton General Hospital, University Hospital Southampton NHS Foundation Trust, Southampton, UK**

David Land (PI), Helen Wheeler, Matt Harvey, Dr Mark Watson, Dr Michael Brown, Dr Ben Irving, Julie Bigg, Mae Felongco.

**Victoria Hospital, Kirkcaldy, NHS Fife, UK**

Dr Joe Mackenzie (PI), Dr Devesh Dhasmana, Dr Rob Thompson, Dr Patrick Lui, Fiona Adam, Fleur Davey, Julie Penman, Amanda McGregor and Patricia Cochrane.

**Walsall Manor Hospital, Walsall Healthcare NHS Trust, Walsall, UK**

Dr Korah Shalan (PI).

**Warwick Clinical Trials Unit, University of Warwick, Coventry, UK**

Will Bozic, Jaclyn Brown, John Carey, Claire Daffern, Emily Dight, Matthew Gane, Belinder Ghuman, Jo Grummett, Johnny Guck, Louisa Hamilton, Cat Hill, Maddy Hill, Chockalingam Muthiah, Emma Padfield, Jeskaran Rai, Kerry Raynes, Greg Scott, Emily Stimpson, Natalie Strickland, Adrian Willis, Jill Wood.

**Warwick Hospital, South Warwickshire NHS Foundation Trust, Warwick, UK**

Dr Ben Attwood (PI), Inderjit Atwal, Penny Parsons.

**Watford General Hospital, West Hertfordshire Hospitals NHS Trust, Watford, UK**

Dr Rama Vancheeswaran (PI), Dr Shruthi Konda, Dr Yadee Maung Maung Myint, Dr Meera Mehta, Dr Ambreen Muhammad, Dr Alessio Navarro, Adam Rochester, Saul Sundayi.

**Wishaw General Hospital, NHS Lanarkshire, Wishaw, UK**

Prof Manish Patel (PI), Prof Andrew Smith, Dr Colin Stewart, Dr Matthew Tate, Dr Erin McGarry, Dr Claire (Rebecca) Pearson, Berni Welsh, Lynn Glass, Karen Black, Suzanne Clements, Rosalind Boyle, Chloe MacDonald, Leigh Hamilton, Gayle Moreland, Raymond Hamill.

**Wrexham Maelor Hospital, Betsi Cadwaladr University Health Board, Wrexham, UK**

Dr Harsha Reddy (PI), Sara Smuts.

**Wythenshawe Hospital, Manchester University NHS Foundation Trust, UK**

Andrew Bentley (PI).

FIGURE S1: RECOVERY RS trial recruitment and UK hospitalized COVID-19 patients

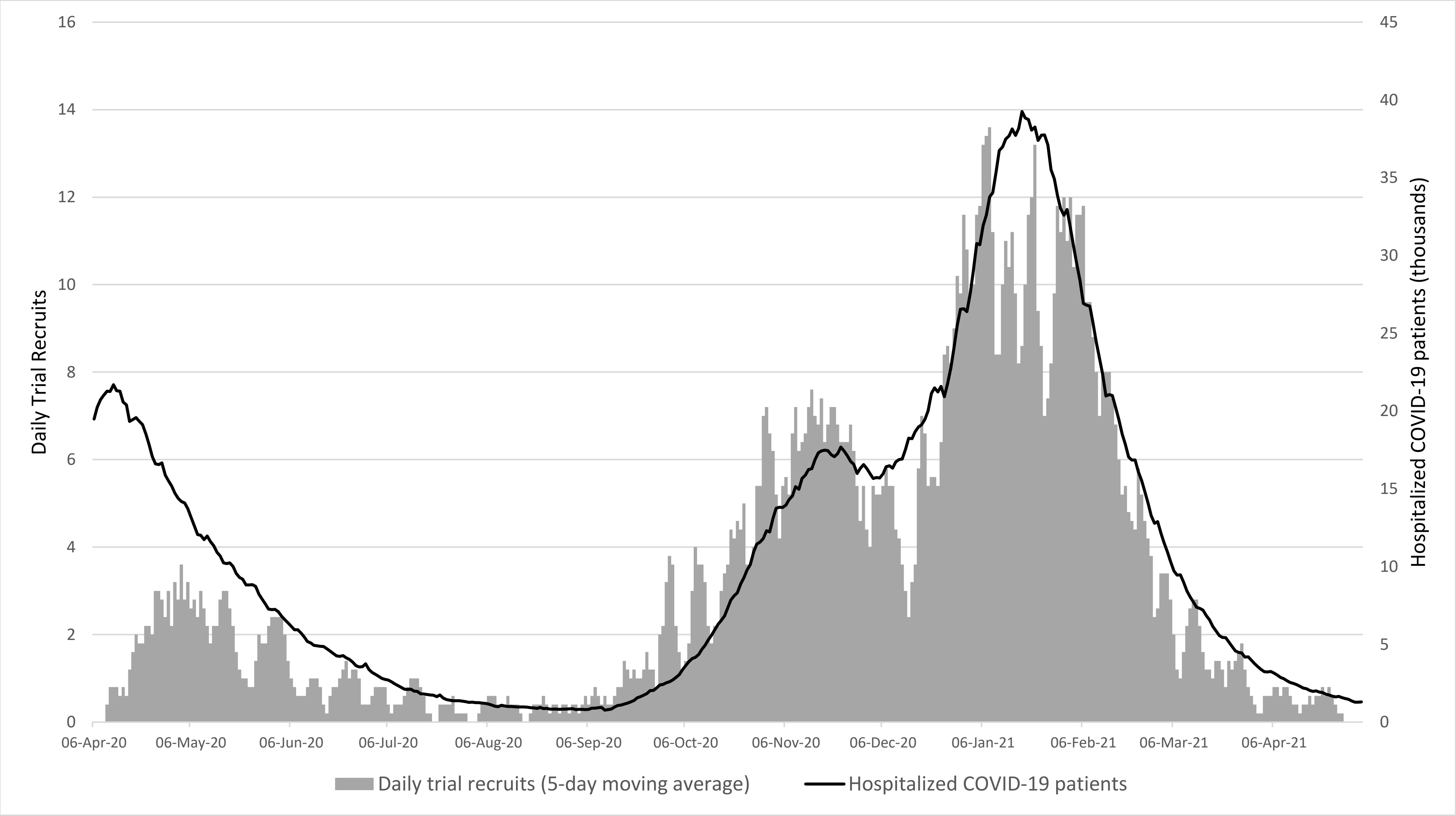


Data for hospitalized COVID-19 patients extracted from publically available UK data sources at https://coronavirus.data.gov.uk/details/healthcare

Table S1: Summary of randomizations and data for pairwise comparisons

| Randomization | Treatment arm | | |
|---|---|---|---|
| | **CONVENTIONAL OXYGEN THERAPY** | **CPAP** | **HFNO** |
| **Device availability** | | | |
| CPAP and conventional oxygen therapy only | 103 | 114 | NA |
| HFNO and conventional oxygen therapy only | 113 | NA | 109 |
| CPAP, HFNO and conventional oxygen therapy | 259 | 266 | 308 |
| Total | 475 | 380 | 417 |
| **Pairwise comparison** | **Treatment arm** | | |
| | **CONVENTIONAL OXYGEN THERAPY** | **CPAP** | **HFNO** |
| CPAP vs CONVENTIONAL OXYGEN THERAPY | 362 (48.8%) | 380 (51.2%) | NA |
| HFNO vs CONVENTIONAL OXYGEN THERAPY | 372 (47.1%) | NA | 417 (52.9%) |

Key: CPAP- Continuous Positive Airway Pressure; HFNO- High-flow nasal oxygen

Table S2: Summary of trial crossover by treatment arm

| Category of crossover | n/N (%) |
|---|---|
| Participants randomized to CPAP | |
| Received HFNO | 58/380 (15.3%) |
| Participants randomized to HFNO | |
| Received CPAP | 48/417 (11.5%) |
| Participants randomized to conventional oxygen therapy | |
| Received CPAP | 40/475 (8.4%) |
| Received HFNO | 36/475 (7.6%) |
| Received both CPAP and HFNO | 36/475 (7.6%) |

Key: CPAP- Continuous Positive Airway Pressure; HFNO- High-flow nasal oxygen

Table S3: Additional participant baseline characteristics

| **Characteristic** | Conventional Oxygen Therapy | CPAP | HFNO | P value |
|---|---|---|---|---|
| Core body temperature at hospital admission | | | | |
| No. of patients | 472 | 377 | 413 | |
| Mean ± SD – (°C) | 37.6 ± 1.0 | 37.7 ± 1.0 | 37.7 ± 1.0 | 0.481* |
| Heart Rate | | | | |
| No. of patients | 469 | 371 | 410 | |
| Mean ± SD – (bpm) | 88.7 ± 16.7 | 91.1 ± 17.0 | 90.4 ± 15.8 | 0.095* |
| Systolic blood pressure | | | | |
| No. of patients | 473 | 377 | 414 | |
| Mean ± SD – (mmHg) | 127.3 ± 18.1 | 128.6 ± 19.0 | 128.4 ± 18.1 | 0.521* |
| Diastolic blood pressure | | | | |
| No. of patients | 472 | 374 | 414 | |
| Mean ± SD – (mmHg) | 75.6 ± 11.2 | 75.2 ± 12.4 | 75.4 ± 12.2 | 0.859* |
| Urea | | | | |
| No. of patients | 464 | 372 | 406 | |
| Mean ± SD – (mg/dl) | 39.4 ± 23.9 | 39.4 ± 23.7 | 41.0 ± 24.9 | 0.542* |
| $PaCO_2$ (partial pressure of carbon dioxide) | | | | |
| No. of patients | 332 | 253 | 305 | |
| Test not available | 139 | 120 | 108 | |
| Mean ± SD (mmHg) | 34.3 ± 6.3 | 33.6 ± 5.3 | 33.4 ± 6.2 | 0.097* |
| Confusion – no. (%) | | | | |
| Confused | 9 (1.9) | 14 (3.7) | 9 (2.2) | 0.614# |
| Not confused | 461 (97.1) | 364 (95.8) | 405 (97.1) | |
| N/A- sedated | 1 (0.2) | 0 | 1 (0.2) | |
| CURB-65 Score – no. (%) | | | | 0.791# |
| 0 | 170 (35.8) | 133 (35.0) | 136 (32.6) | |
| 1 | 176 (37.1) | 129 (34.0) | 148 (35.5) | |
| 2 | 88 (18.5) | 71 (18.7) | 85 (20.4) | |
| 3 | 22 (4.6) | 30 (7.9) | 30 (7.2) | |
| 4 | 3 (0.6) | 2 (0.5) | 3 (0.7) | |
| 5 | 1 (0.2) | 0 (0.0) | 0 (0.0) | |
| Treatment phases – no. (%) | | | | 0.798# |
| Before July 2020 | 47 (9.9) | 47 (12.4) | 44 (10.6) | |
| July 2020 - January 2021 | 331 (69.7) | 262 (69.0) | 288 (69.1) | |
| After January 2021 | 97 (20.4) | 71 (18.7) | 85 (20.4) | |
| CPAP device type – no. (%) | | | | <0.001# |
| NIV device in CPAP mode | 47 (9.9) | 147 (38.7) | 37 (8.9) | |
| CPAP device | 35 (7.4) | 173 (45.5) | 25 (6.0) | |
| Other | 10 (2.1) | 24 (6.3) | 8 (1.9) | |

Key- CPAP- Continuous Positive Airway Pressure; HFNO- High-flow nasal oxygen; N/A- Not applicable; NIV- Non-invasive ventilation
* T-test used to calculate p-value; # Chi-squared statistics used to calculate the p-value.

Table S4: Adverse events and serious adverse events by treatment arm

| | Conventional oxygen therapy (n=475) | CPAP (n=380) | HFNO (n=417) | P-value‖ | All participants (n=1272) |
|---|---|---|---|---|---|
| Participants with AE/ SAE- n (%) | 66 (13.9%) | 130 (34.2%) | 86 (20.6%) | <0.001 | 282 (22.2%) |
| **ADVERSE EVENTS** | | | | | |
| **Participants with AE- n (%)** | 65 (13.7%) | 130 (34.2%) | 86 (20.6%) | <0.001 | 281 (22.1%) |
| **Summary of events- n(%)*** | | | | | |
| Interface/therapy Intolerance | 1 (0.2%) | 22 (5.8%) | 3 (0.7%) | | 26 (2.0%) |
| Pain | 26 (5.5%) | 13 (3.4%) | 13 (3.1%) | - | 52 (4.1%) |
| Cutaneous pressure sore | 14 (2.9%) | 32 (8.4%) | 14 (3.4%) | - | 60 (4.7%) |
| Claustrophobia | 28 (5.9%) | 11 (2.9%) | 16 (3.8%) | - | 55 (4.3%) |
| Oronasal dryness | 9 (1.9%) | 25 (6.6%) | 25 (6.0%) | - | 59 (4.6%) |
| Respiratory acidosis | 4 (0.8%) | 4 (1.1%) | 11 (2.6%) | - | 19 (1.5%) |
| Haemodynamic instability | 29 (6.1%) | 43 (11.3%) | 36 (8.6%) | - | 108 (8.5%) |
| Nausea and vomiting | 6 (1.3%) | 9 (2.4%) | 10 (2.4%) | - | 25 (2.0%) |
| Aspiration of gastric contents | 2 (0.4%) | 6 (1.6%) | 5 (1.2%) | - | 13 (1.0%) |
| Pneumothorax | 11 (2.3%) | 7 (1.8%) | 8 (1.9%) | - | 26 (2.0%) |
| Pneumomediastinum | 5 (1.1%) | 12 (3.2%) | 3 (0.7%) | - | 20 (1.6%) |
| Anxiety and confusion | 0 | 6 (1.6%) | 3 (0.7%) | - | 9 (0.7%) |
| Pulmonary embolism | 1 (0.2%) | 1 (0.3%) | 0 | - | 2 (0.2%) |
| Surgical emphysema | 0 | 3 (0.8%) | 1 (0.2%) | - | 4 (0.3%) |
| Haemoptysis | 0 | 1 (0.3%) | 1 (0.2%) | - | 2 (0.2%) |
| Other† | 1 (0.2%) | 5 (1.3%) | 8 (1.9%) | - | 14 (1.1%) |
| **SERIOUS ADVERSE EVENTS** | | | | | |
| **Participants with SAE- n (%)‡** | 1 (0.2%) | 7 (1.8%) | 0 (0.0%) | 0.002 | 8 (0.6%) |
| **Impact of SAE*** | | | | | |
| Death | 1 (0.2%) | 1 (0.3%) | 0 (0.0%) | | 2 (0.2%) |
| Life Threatening | 0 (0.0%) | 4 (1.1%) | 0 (0.0%) | | 4 (0.3%) |
| Hospitalisation | 0 (0.0%) | 6 (1.6%) | 0 (0.0%) | | 6 (0.5%) |
| Disability | 0 (0.0%) | 1 (0.3%) | 0 (0.0%) | | 1 (0.1%) |
| Birth Defect | 0 (0.0%) | 0 (0.0%) | 0 (0.0%) | | 0 (0.0%) |
| Required Intervention | 0 (0.0%) | 4 (1.1%) | 0 (0.0%) | | 4 (0.3%) |
| **Causality of SAE** | | | | | |
| Definitely | 0 (0.0%) | 0 (0.0%) | 0 (0.0%) | | 0 (0.0%) |
| Probably | 0 (0.0%) | 1 (0.3%) | 0 (0.0%) | | 1 (0.1%) |
| Possibly | 0 (0.0%) | 3 (0.8%) | 0 (0.0%) | | 3 (0.2%) |
| Unlikely | 0 (0.0%) | 2 (0.5%) | 0 (0.0%) | | 2 (0.2%) |
| Unrelated | 1 (0.2%) | 1 (0.3%) | 0 (0.0%) | | 2 (0.2%) |

Key- AE- Adverse event; CPAP- Continuous Positive Airway Pressure; HFNO- High-flow nasal oxygen; SAE- Serious adverse event
‖- p-value calculated using chi-square test for AE/SAE and AE comparison, and using Fisher-exact for SAE comparison
*Multiple events/categories allowed per participants
†Details of other events:
Conventional oxygen therapy (one event): Nasal cannulae leak
CPAP (five events): Chest tightness; Significant desaturation when eating; CPAP leak; Pneumopericardium; Low tidal volume/hypoxia/dyspnoea (one of each event)
HFNO (eight events): Abdominal distension; Bilateral rupture of tympanic membrane; Monoclonal antibody treatment side-effect (hand pustules); Need for tracheostomy; Ventilator-associated pneumonia and klebsiella meningitis diagnosis; Pleural

effusions; Secondary sepsis, intracranial bleed, requirement for renal replacement therapy; Detail not reported (one of each event)

‡Details of serious adverse events:
Conventional oxygen therapy (one event): Pulmonary embolus
CPAP (seven events): Type 2 myocardial infarction (one event); surgical emphysema and pneumomediastinum (one event); vomiting requiring emergency tracheal intubation (one event); Intracranial bleed (one event); Perforated bowel (one event); Pneumothorax and pneumomediastinum (two events)
